# DunedinPACE Predicts Incident Metabolic Syndrome: Cross-sectional and Longitudinal Data from the Berlin Aging Study II (BASE-II)

**DOI:** 10.1101/2024.12.16.24319083

**Authors:** Ilja Demuth, Valentin Max Vetter, Jan Homann, Vera Regitz-Zagrosek, Denis Gerstorf, Christina M. Lill, Lars Bertram

## Abstract

**Importance:** Data on the capacity of more recently developed epigenetic age measures to predict a future onset of Metabolic Syndrome (MetS) are lacking.

**Objective:** The aim of the study was a comparative analysis of different DNA methylation (DNAm)-based epigenetic clocks with regard to their ability to predict a future onset of MetS. In addition, cross-sectional relationships between epigenetic age measures, MetS and its components were investigated.

**Design, Setting and Participants:** MetS was diagnosed in participants of the Berlin Aging Study II at baseline (n=1,671, mean age 68.8 ±3.7 years, 51.6% women) and at follow-up (n=1,083; 7.4 ±1.5 years later). DNAm age (DNAmA) and its deviation from chronological age, i.e., DNAmA acceleration (DNAmAA), were calculated for a total of five epigenetic clocks at baseline. In addition, DunedinPACE, a DNAm-based measure of the pace of aging, was calculated. The relationship of MetS with DNAmAA and DunedinPACE was investigated by fitting regression models. Furthermore, receiver operating characteristic statistics were calculated to investigate the capacity of DNAm clocks assessed at baseline to predict incident MetS at follow-up.

**Exposures:** Six different epigenetic age measures including DunedinPACE assessed at baseline to predict MetS in the future.

**Main Outcomes and Measures:** Diagnosis of incident MetS on average 7.4 ±1.5 years after baseline.

**Results:** DunedinPACE was associated with incident MetS at follow-up on average 7.4 years later (OR: 9.84, p=0.028). Interestingly, we observed no significant differences (p>0.05) in the area under the curve in predicting MetS between a model that only included clinical parameters and a model that only used GrimAge DNAmAA. Cross-sectional differences between participants with and without MetS remained statistically significant for DunedinPACE only after covariate adjustment (baseline: β=0.042, follow-up: β=0.031, p<0.0001 in both cases).

**Conclusions and Relevance:** Systematic comparison of epigenetic clocks within a single dataset in relation to MetS and its diagnostic components showed strong and consistent associations with DunedinPACE, but not with other epigenetic clocks. Our results highlight the potential of using certain DNAm-based measures of biological ageing in predicting the onset of clinical outcomes, such as MetS.

**Key Points:** *Question:* Are epigenetic age measures able to predict the future onset of Metabolic Syndrome?

*Findings:* This longitudinal observational study revealed that individuals with a one-year faster pace of aging (DunedinPACE) had 2.3-fold increased odds for incident Metabolic Syndrome ∼7.5-years later.

*Meaning:* The data reported here will potentially help to implement epigenetic measures in clinical risk assessment.

## Introduction

Metabolic Syndrome (MetS) is defined as a cluster of cardiovascular (CV) and type 2 diabetes (T2D) risk factors, which frequently occur together ^1^. MetS is diagnosed when at least three of the following five metabolic phenotypes are present: hypertension, dyslipidemia (hypertriglyceridemia and/or low high-density lipoprotein cholesterol (HDL-C)), hyperglycemia, and abdominal obesity. With a worldwide prevalence of up to 30% among adults and similar numbers in Europe and Germany ^2,3^, MetS is of significant public health relevance. The relationship between MetS and various biomarkers of aging has been investigated and respective associations have been reported for biomarkers such as telomere length, epigenetic, transcriptomic and proteomic clocks (e.g. ^4–6^). Given the abovementioned clinical implications of MetS, this type of research is not only of academic interest, but also of high translational relevance e.g. in the context of being able to predict MetS incidence in the future.

Epigenetic aging measures, also known as epigenetic clocks, were developed during the past decade and are believed to reflect estimators of “biological age”. As such, they have been reported to show associations with a large number of factors linked to morbidity and mortality (reviewed e.g. in ^7,8^). While the so-called first-generation clocks (e.g. Horvath and Hannum) were trained to predict chronological age, clocks of the second (e.g. PhenoAge, GrimAge) and third (e.g. DunedinPACE) generation were developed using a range of phenotypic measures (e.g. laboratory parameters and lifestyle factors) and mortality as training variables. One typical variable of interest is the estimation of the degree of DNAmA acceleration (DNAmAA), which can be quantified via the residual of a linear regression of DNAmA on chronological age. DNAmAA reflects the degree of accelerated (values >0) or decelerated (values <0) aging in years.

Although epigenetic clocks have been the topic of intensive research over the past years, to date only few studies report on the cross-sectional relationship between diagnosed MetS and DNAmAA. These studies largely agree that DNAmAA as estimated by the GrimAge algorithm is associated with MetS cross-sectionally ^9–11^ but not with DNAmAA calculated from another second-generation clock, PhenoAge, or the first-generation Hannum clock ^9^. Only one study investigated MetS cross-sectionally using the DunedinPACE aglorithm ^11^, a third-generation clock. This clock was trained to predict a longitudinal biological aging measure, Pace of Aging, aggregated from data on a variety of age-associated phenotypic markers assessed at four time points over 20 years in the Dunedin study ^12^. We are only aware of one study using this marker showing that MetS was cross-sectionally associated with GrimAge DNAmAA and DunedinPACE after minimal covariate adjustment ^11^.

Longitudinal data on epigenetic age measures in the context of MetS are even more scarce. To our knowledge, only Nannini and colleagues investigated this relationship longitudinally ^6^. They reported evidence for an association between Horvath (but not Hannum)-derived DNAmAA and incident MetS diagnosed 10 years later. No second- or third-generation clocks were analyzed in this study.

In the current study we assessed the relationship between MetS and its components and epigenetic age measures derived from DNAm clock algorithms from all three generations in up to n=1,021 participants of the Berlin Aging Study II (BASE-II). In addition to cross-sectional analyses, we also investigated these relationships longitudinally over a mean follow-up period of on average about 7.5 years. Our work contributes to closing the knowledge gap on the role of more recently developed epigenetic age measures in MetS as well as on the clocks’ predictive value for a future onset of MetS.

## Methods

### Berlin Aging Study II (BASE-II)

BASE-II is an observational, longitudinal, and multi-disciplinary study that investigates factors promoting “healthy” vs. “unhealthy” aging. At baseline (2009-2014), the medical part of BASE-II included 1,671 participants of the greater Berlin metropolitan area between the age of 60-85 years (older group) ^13–15^. Men and women were recruited in equal numbers (see also Supplementary Methods). A comprehensive medical follow-up including 1,083 participants of the older group took place between 2018 and 2020 after an average follow-up period of 7.4 (±1.5 SD) years ^16^.

All participants gave written informed consent. The medical assessments at baseline and follow-up were conducted in accordance with the Declaration of Helsinki and approved by the Ethics Committee of the Charité – UniversitaLJtsmedizin Berlin (approval numbers EA2/029/09 and EA2/144/16).

### Estimators of epigenetic age and DunedinPACE

DNAm age according to Horvath’s clock ^17^, Hannum’s clock ^18^, PhenoAge ^19^, GrimAge ^20^, and DunedinPACE ^21^ were estimated from DNAm data measured by Illumina’s “Infinium MethylationEPIC” array (Illumina Inc., USA). Details on DNA processing, QC and sample processing are described elsewhere ^22^ (see also Supplementary Methods). In addition, we also estimated DNAmA using the “7-CpG clock” ^23^ (see also Supplementary Methods).

### DNA methylation age acceleration

To adjust for known age-associated changes in leukocyte cell composition, we calculated DNAmAA as residuals from a cell-count (neutrophils, monocytes, lymphocytes, eosinophils)-adjusted linear regression analysis of DNAmA on chronological age ^23,24^. All cell counts were determined in a certified laboratory by flow cytometry (MVZ Labor 28 GmbH, Berlin, Germany).

### Metabolic Syndrome (MetS) and its components

MetS was diagnosed according to Alberti et al. ^1^ when three or more of the following five components were present:

– *Abdominal obesity:* waist circumference ≥94 cm in men and ≥80 cm in women
– *High triglycerides:* triglycerides ≥150 mg/dl
– *Low HDL-C:* HDL-C <40 mg/dl in men; <50 mg/dl in women
– *High blood pressure:* systolic blood pressure ≥130 and/or diastolic blood pressure ≥85 mmHg or prevalent hypertension or use of antihypertensive medication in participants with known hypertension
– *Insulin resistance:* fasting glucose ≥100 mg/dL, prevalent T2D or use of antidiabetic medication in participants with T2D.

### Covariates

Sex differences in the aging process and between epigenetic aging-markers are well documented ^25–27^, therefore sex was included as a covariate. Furthermore, we performed sex-stratified analyses whose results are presented in Tables S2-S6. Overall morbidity was assessed using a modified version ^28^ of Charlson’s co-morbidity index ^29^. Additionally, smoking behavior (packyears), alcohol consumption (g/d) assessed via a food frequency questionnaire ^30^, and physical activity (answer to the question “Are you seldom or never physically active” (yes/no)) were considered.

### Simple model for predicting metabolic syndrome

A clinical model guided by variables described by Tan et. al. ^31^ including Hemoglobin A1c (HbA1c), systolic blood pressure (BP), waist-to-hip ratio (WHR), age and sex was calculated to compare its performance to models including either additionally GrimAge DNAmAA or DunedinPACE or the latter as single MetS predictors (see also Supplementary Methods).

### Statistical Analyses

Characteristics of the study population are provided as mean (standard deviation, SD), median (interquartile range, IQR) or as numbers and percentages, as appropriate. Group comparisons (one-way analyses of variance (ANOVA), t-tests, Mann-Whitney-U-Tests or χ2 tests) and calculations of Cohen’s d were performed using the SPSS IBM Software package, version 29. Column scatter plots were generated using the GraphPad Prism software version 6 for Windows (GraphPad Software, Boston, Massachusetts USA). Linear and logistic regression analyses were calculated using lm- and glm-function in R (version 4.4.1) ^32^. ROC statistics were calculated and AUC curves were created using the pROC-package. ROC curves were compared using the roc.test function from the pROC package. Statistical significance was defined at alpha=0.05.

## Results

### Characteristics of BASE-II participants at baseline and follow-up

For the cross-sectional analyses, we used BASE-II data from all participants of the older age group (i.e. ≥60 years at recruitment), for which all DNAm clocks were available at baseline and the MetS status was known at baseline and follow-up. This resulted in an effective sample size of n=902 participants (50.2% women) aged 68.2 (±3.5) years (Table 1). With the exception of blood pressure, all parameters used for the diagnosis of the MetS differed in statistically significant ways between participants with and without MetS. This was also observed for the MetS risk factors, including high BMI, physical inactivity, and smoking (Table 1). MetS prevalence was higher in men compared to women (41.2% vs. 27.6%, overall prevalence: 34.4%).

**Table 1:** Descriptive statistics of participants at baseline stratified for Metabolic Syndrome (MetS).

|  | n | All<br>(n=902) | No MetS<br>(n=592) | MetS<br>(n=310) | p-value |
| --- | --- | --- | --- | --- | --- |
| Chronological age (y) | 902 | 68.2 (3.5) | 68.3 (3.4) | 68.1 (3.7) | 0.407 |
| Sex (female) (%) | 902 | 453 (50.2) | 328 (55.4) | 125 (40.3) | <0.001 |
| Morbidity Index<sup>1</sup> | 844 | 1 (2) | 0 (1) | 1 (2) | <0.001 |
| BMI (kg/m<sup>2</sup>) | 899 | 26.6 (4.0) | 25.4 (3.5) | 29 (3.9) | <0.001 |
| Waist (cm) | 902 | 95.4 (11.6) | 91.7 (10.7) | 102.3 (10.0) | <0.001 |
| Systolic BP (mmHg) | 902 | 147.8 (48.7) | 146.2 (50.3) | 150.8 (45.5) | 0.181 |
| Diastolic BP (mmHg) | 902 | 88.2 (48.8) | 87.8 (50.1) | 88.7 (46.3) | 0.798 |
| Triglycerides (mg/dL) | 902 | 111.7 (56.1) | 91.3 (31.1) | 150.7 (70.7) | <0.001 |
| Fasting Glucose (mg/dL) | 896 | 94.8 (17.6) | 88.5 (8.9) | 106.9 (23.0) | <0.001 |
| LDL-C (mg/dL) | 902 | 130.2 (34.7) | 131.4 (33.9) | 127.8 (36.2) | 0.134 |
| HDL-C (mg/dL) | 902 | 62.3 (16.8) | 67.0 (15.5) | 53.5 (15.7) | <0.001 |
| Physically Inactive (%) | 900 | 73 (8.1) | 34 (5.8) | 39 (12.6) | <0.001 |
| Smoking (packyears)<sup>1</sup> | 867 | 0 (14) | 0 (10) | 3 (20) | <0.001 |
| Alcohol Intake (g/d)<sup>1</sup> | 890 | 9.1 (17) | 8.8 (16) | 9.6 (19) | 0.07 |
| Number of MetS Components | 902 |  |  |  |  |
| 0 |  |  | 36 |  |  |
| 1 |  |  | 181 |  |  |
| 2 |  |  | 375 |  |  |
| 3 |  |  |  | 198 |  |
| 4 |  |  |  | 87 |  |
| 5 |  |  |  | 25 |  |
| Horvath DNAmA (y) | 902 | 63.8 (4.9) | 63.5 (4.7) | 64.3 (5.1) | 0.012 |
| Horvath DNAmAA (y) | 902 | 0.07 (4.23) | -0.20 (4.13) | 0.59 (4.36) | 0.07 |
| GrimAge DNAmA (y) | 902 | 75.2 (4.3) | 74.8 (4.2) | 76.0 (4.4) | <0.001 |
| GrimAge DNAmAA (y) | 902 | 0.03 (0.04) | -0.32 (2.93) | 0.71 (3.13) | <0.001 |
| DunedinPACE (y) | 902 | 1.03 (0.11) | 1.01 (0.11) | 1.06 (0.11) | <0.001 |

BMI: body mass index, MetS: metabolic syndrome, BP: blood pressure, LDL-C: low density lipoprotein-cholesterol, HDL-C: high density lipoprotein-cholesterol, DNAmA: DNA methylation age, DNAmAA: DNA methylation age acceleration. Unless otherwise indicated, data are given as mean ± standard deviation or as figures and percentages in brackets. <sup>1</sup>Data are shown as median and interquartile range.

In addition to analyses in the baseline sample, we also tested for differences at the time of follow-up (n=1,021; 75.6 ±3.7; 47.8% women, Supplementary Table S1). Compared to baseline, the follow-up dataset was slightly larger due to fewer missing values. At follow-up, MetS was diagnosed in 45.3% of the participants of which 55.3% were men (Supplementary Table S1). The differences in the individual MetS components and MetS-associated variables between the participants with and without MetS were similar to the differences we found at baseline (Supplementary Table S1).

### Accelerated epigenetic aging in participants with MetS in cross-sectional analyses

Comparison of epigenetic age estimates between participants with and without MetS at baseline revealed statistically significant differences with higher Horvath and GrimAge DNAmAA and higher pace of aging (DunedinPACE) in the MetS group (t-tests, p<0.001, Figure 1). At follow-up, similar differences were observed for GrimAge and DunedinPACE, but not for the Horvath DNAmAA estimate (Figure 1). At both timepoints, we found a gradual increase in effect sizes with higher DNAm clock generations. The lowest effect size was detected for the Horvath clock (baseline: d=0.19; follow-up: d=0.12) and the highest effect size was detected for DunedinPACE (baseline: d=0.48; follow-up: d=0.40).

**Figure 1:**
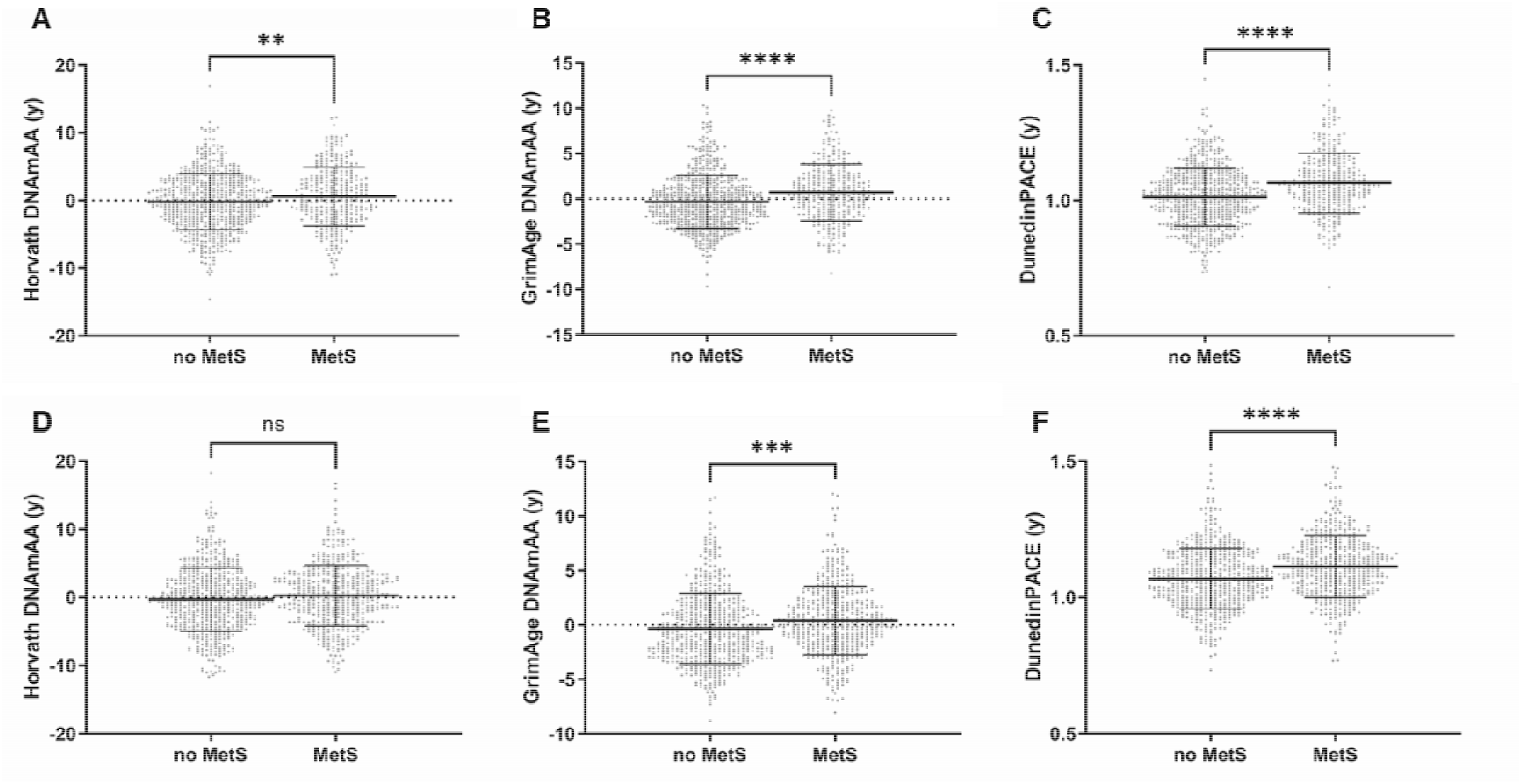
Comparison of epigenetic clocks in BASE-II participants with and without MetS. Column scatter plots comparing DNAmAA or the pace of aging estimated by the Horvath and GrimAge clocks (DNAmAA) and DunedinPACE as assessed at baseline (A-C) and at follow- up (D-F) between participants diagnosed with MetS and participants without MetS. All participants of the older age group with data on MetS and all biological age estimators available are included (baseline: n=902. follow-up: n=1,021). Mean and standard deviation is indicated and the p-values were determined by t-tests. DNAmAA = DNA methylation age acceleration. **p<0.01, ***p<0.001, ****p<0.0001.

To further elucidate the identified associations, we investigated the epigenetic markers in relation to the number of MetS diagnostic criteria that were met by the BASE-II participants. These analyses revealed that with an increasing number of MetS criteria fulfilled, there was a corresponding increase in DNAmAA and DunedinPACE. The associations between the number of individual MetS components and epigenetic biomarkers were statistically significant at both time points for GrimAge DNAmAA and DunedinPACE (ANOVA, p<0.001, Figure 2), but not for the Horvath clock.

**Figure 2:**
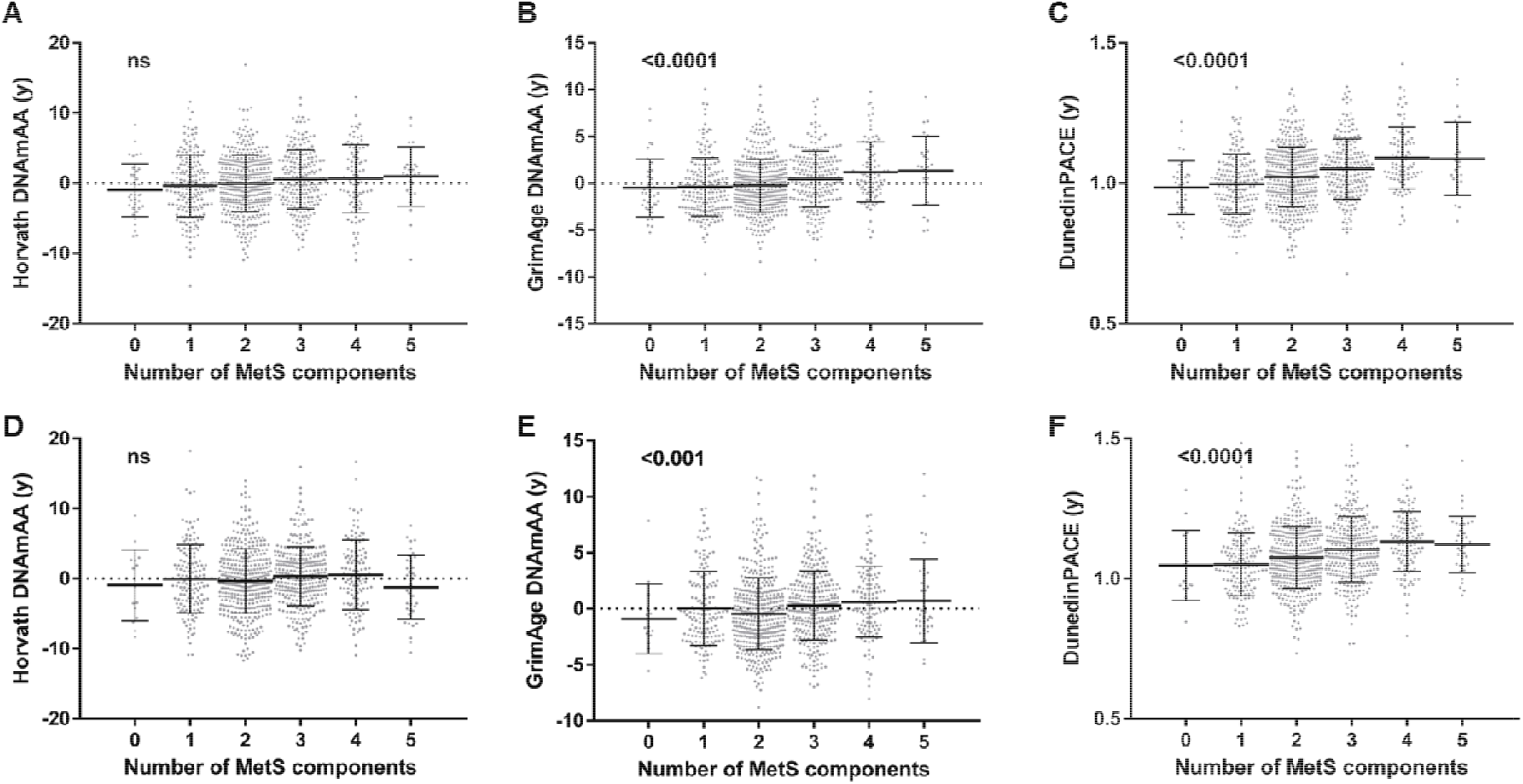
Comparison of epigenetic clocks in BASE-II participants in relation to the number of MetS components fulfilled. Column scatter plots of the number of MetS components and biological age estimators at baseline (A-C) and follow-up (D-F). All participants of the older age group with data on MetS and all biological age estimators are included (baseline: n=902, follow-up: n=1,021). Mean and standard deviation is indicated and the p-values were determined by one-way ANOVA. DNAmAA = DNA methylation age acceleration. **p<0.01, ***p<0.001, ****p<0.0001.

After adjustment for covariates (chronological age, sex, overall morbidity, smoking, physical activity, and alcohol consumption) DunedinPACE remained significantly higher in participants with MetS compared to participants without MetS at baseline and follow-up (β=0.042 and β=0.031, p<0.0001; Table 2; Supplementary Table 2 and 3). Similarly, the association between DunedinPACE and the number of MetS categories fulfilled remained statistically significant in the fully adjusted model at both time points (model 3, β=0.02 and β=0.016, p<0.0001, Tables 3, Supplementary Tables S4 and S5). In addition, we found the Horvath DNAmAA to be statistically significantly associated with the MetS categories at baseline in the fully adjusted model (β=0.294, p=0.037).

**Table 2:** Linear regression analysis of epigenetic age estimators on MetS status at baseline.

| Dependent Variable | Model<sup>1</sup> | $\beta$ | SE | p | lowCI | upCI | n |
| --- | --- | --- | --- | --- | --- | --- | --- |
| Horvath Clock | 1 | 0.602 | 0.296 | 0.042 | 0.022 | 1.183 | 902 |
|  | 2 | 0.553 | 0.308 | 0.073 | -0.052 | 1.159 | 844 |
|  | 3 | 0.476 | 0.324 | 0.141 | -0.159 | 1.112 | 801 |
| GrimAge | 1 | 0.651 | 0.194 | 0.001 | 0.270 | 1.033 | 902 |
|  | 2 | 0.727 | 0.204 | <0.001 | 0.328 | 1.127 | 844 |
|  | 3 | 0.352 | 0.194 | 0.070 | -0.029 | 0.734 | 801 |
| DunedinPACE | 1 | 0.046 | 0.007 | <0.0001 | 0.032 | 0.061 | 902 |
|  | 2 | 0.048 | 0.008 | <0.0001 | 0.033 | 0.063 | 844 |
|  | 3 | 0.042 | 0.008 | <0.0001 | 0.026 | 0.058 | 801 |

<sup>1</sup>Model 1 is adjusted for age and sex. Model 2 is additionally adjusted for overall morbidity and model 3 includes lifestyle factors (smoking (packyears), physical activity and alcohol consumption (g/d) in addition to the covariables considered in model 2. St. = Standardized; SE = Standard Error; p = p-value; lowCI = lower 95% confidence interval; upCI = upper 95% confidence interval.

**Table 3:** Linear regression analysis of epigenetic age estimators on the number of MetS categories fulfilled at baseline.

| Dependent Variable | Model<sup>1</sup> | $\beta$ | SE | p | lowCI | upCI | n |
| --- | --- | --- | --- | --- | --- | --- | --- |
| Horvath Clock | 1 | 0.322 | 0.129 | 0.013 | 0.068 | 0.576 | 902 |
|  | 2 | 0.308 | 0.133 | 0.020 | 0.048 | 0.569 | 844 |
|  | 3 | 0.294 | 0.140 | 0.037 | 0.018 | 0.569 | 801 |
| GrimAge | 1 | 0.326 | 0.085 | <0.001 | 0.159 | 0.493 | 902 |
|  | 2 | 0.334 | 0.088 | <0.001 | 0.162 | 0.506 | 844 |
|  | 3 | 0.150 | 0.084 | 0.075 | -0.015 | 0.316 | 801 |
| DunedinPACE | 1 | 0.024 | 0.003 | <0.0001 | 0.017 | 0.030 | 902 |
|  | 2 | 0.023 | 0.003 | <0.0001 | 0.017 | 0.030 | 844 |
|  | 3 | 0.020 | 0.003 | <0.0001 | 0.013 | 0.027 | 801 |

<sup>1</sup>Model 1 is adjusted for age and sex. Model 2 is additionally adjusted for overall morbidity and model 3 includes lifestyle factors (smoking (packyears), physical activity and alcohol consumption (g/d) in addition to the covariables considered in model 2. St. = Standardized; SE = Standard Error; p = p-value; lowCI = lower 95% confidence interval; upCI = upper 95% confidence interval.

### DunedinPACE is associated with incident MetS

In a next step, we analyzed if the baseline epigenetic age measures were associated with incident MetS at follow-up (i.e. 7.4 years later). As illustrated in Figure 3, we found a significantly higher GrimAge DNAmAA (p<0.001) and DunedinPACE (p<0.01) at baseline in 141 participants with incident MetS at follow-up compared to those without MetS at follow-up (note that 310 participants were excluded from these analyses due to their MetS diagnosis at baseline). The association between DunedinPACE (baseline) and incident MetS remained statistically significant in logistic regression analyses following full baseline covariate adjustment (Table 4, model 3, OR=2.3, CI 1.3-77.5, p=0.028). Interestingly, none of the other investigated epigenetic age estimates was associated with incident MetS (Table 4 and Supplementary Table S6).

**Figure 3:**
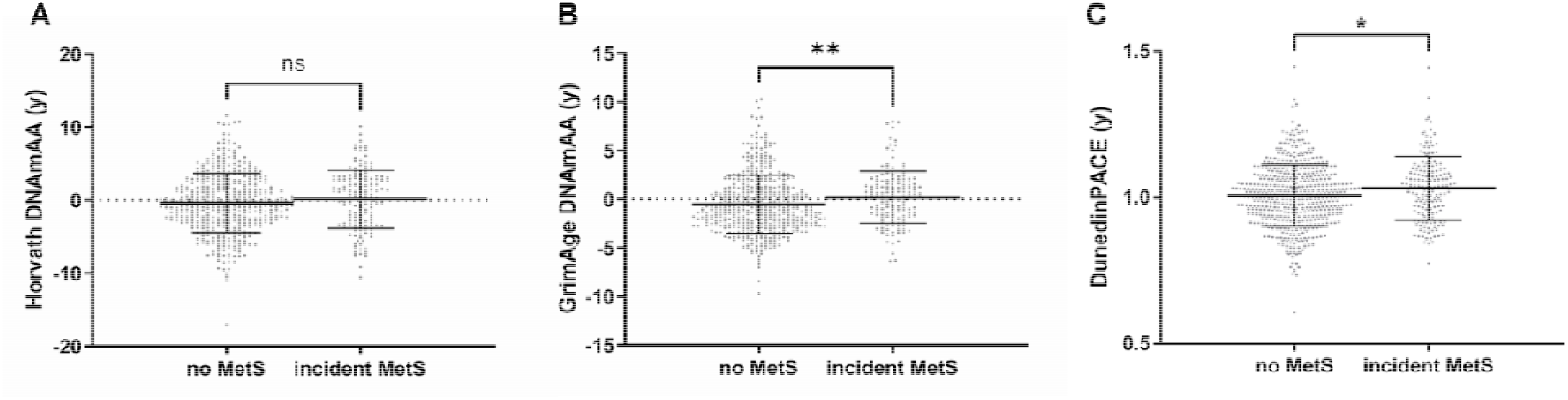
Comparison of epigenetic clocks (baseline) in BASE-II participants with and without incident MetS. Column scatter plots comparing DNAmAA or the pace of aging estimated by the Horvath and GrimAge clocks (DNAmAA) and DunedinPACE as assessed at baseline between participants newly diagnosed with MetS at follow-up (incident MetS) and participants without MetS. All available participants of the older age group with data on MetS and the respective biological age estimator were included. Mean and standard deviation is indicated and the p-values were determined by t-tests. DNAmAA = DNA methylation age acceleration. *p<0.05, **p<0.01.

**Table 4:** Logistic regression analysis of MetS status (incident cases) at follow-up on epigenetic age estimators as assessed at baseline.

| Dependent Variable | Model<sup>1</sup> | $\beta$ | OR | OR lowCI | OR upCI | p | AUC | n |
| --- | --- | --- | --- | --- | --- | --- | --- | --- |
| Horvath Clock | 1 | 0.020 | 1.020 | 0.972 | 1.071 | 0.413 | 0.568 | 575 |
|  | 2 | 0.019 | 1.020 | 0.969 | 1.073 | 0.452 | 0.568 | 542 |
|  | 3 | 0.031 | 1.032 | 0.980 | 1.087 | 0.232 | 0.577 | 513 |
| GrimAge | 1 | 0.052 | 1.054 | 0.983 | 1.129 | 0.140 | 0.582 | 575 |
|  | 2 | 0.061 | 1.063 | 0.989 | 1.143 | 0.096 | 0.590 | 542 |
|  | 3 | 0.071 | 1.073 | 0.986 | 1.168 | 0.100 | 0.585 | 513 |
| DunedinPACE | 1 | 2.085 | 8.048 | 1.266 | 51.934 | 0.027 | 0.587 | 575 |
|  | 2 | 1.861 | 6.431 | 0.946 | 44.236 | 0.057 | 0.589 | 542 |
|  | 3 | 2.286 | 9.835 | 1.287 | 77.457 | 0.028 | 0.601 | 513 |

<sup>1</sup>Model 1 is adjusted for age and sex. Model 2 is additionally adjusted for overall morbidity and model 3 includes lifestyle factors (smoking (packyears), physical activity and alcohol consumption (g/d) in addition to the covariables considered in model 2. All covariates as assessed at baseline. St. = Standardized; SE = Standard Error; p = p-value; lowCI = lower 95% confidence interval; upCI = upper 95% confidence interval.

### DNAmAA and DunedinPACE predict incident MetS 7.4 years later

Lastly, we investigated the capacity of the epigenetic age measures assessed at baseline to predict a future onset of MetS. To this end, we investigated how each of the epigenetic age measures improved a clinical prediction model containing WHR, systolic blood pressure and HbA1c, age, and sex. As can be seen in Figure 4, none of the investigated epigenetic age measures substantially improved the clinical model in discriminating participants with and without incident MetS (all areas under the curves (AUCs) 0.64 or 0.65 without statistically significant differences between analyses, Figure 4 and Supplementary Figure 1). Interestingly, the prediction model including GrimAge DNAmAA as a single predictor reached an AUC that, although slightly smaller, did not differ significantly from the AUC obtained in the model considering only the clinical parameters (Figure 4 and Supplementary Figure 1).

**Figure 4:**
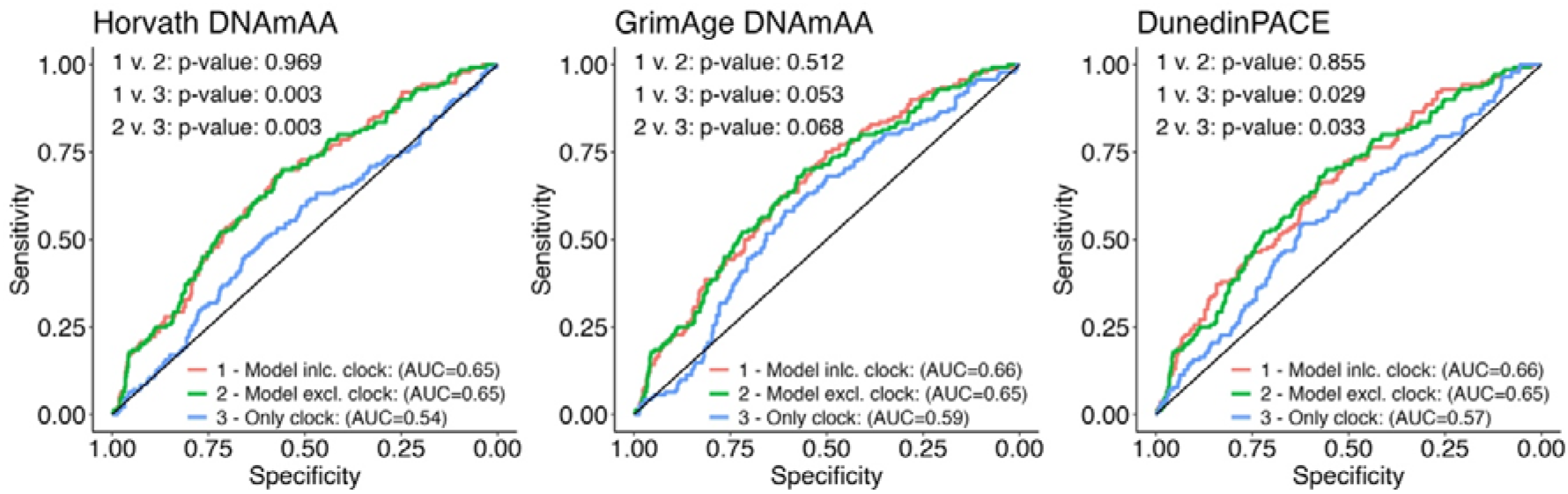
ROC curves derived from logistic regression analyses of incident MetS at follow-up on epigenetic age estimators at baseline. The red (1) and green (2) curve represent prediction model with and without the epigenetic age estimate. The blue curve (3) visualizes results from a prediction model with only the epigenetic age estimate without any additional variables. The blue curve represents a prediction model that includes only the epigenetic estimate. Note: AUC = Area under the curve, DNAmAA = DNA methylation age acceleration; incl.=including; excl.=excluding.

## Discussion

In this study, we investigated the relationship between MetS and epigenetic age measures estimated using DNAm clock algorithms. The main result of these analyses is that baseline DunedinPACE pace of aging estimates are significantly and consistently associated with incident MetS ∼7.4 years later. Individuals with a one-year faster pace of aging had 2.3-fold increased odds to develop incident MetS at follow-up. In general, our study confirmed previous observations of a gradual improvement in the cross-sectional and longitudinal associations of age-associated phenotypes with each additional generation of epigenetic clocks (e.g. ^12,33^). For example, Lee and Park investigated four first- and second-generation clocks in a cohort of n=349 study participants from the Korean Genome Epidemiology Study. This revealed an association between MetS and GrimAge DNAmAA in the subgroup of participants below 60 years of age (Odds Ratio: 1.16) ^9^.

In regression models adjusted for covariates, McCarthy and colleagues found that MetS was associated with about 1 year DNAmAA using GrimAge in a cohort of the Irish Longitudinal Study on Ageing ^10^. Similarly, Föhr and colleagues found 2.6 years higher GrimAge DNAmAA and a 0.12 years faster pace of aging using DunedinPACE in participants with MetS compared to the controls in analyses adjusted for a minimal number of covariates in a cohort of n=268 participants ^11^.

To our knowledge, only one other study investigated the relationship between first-generation epigenetic clocks and MetS longitudinally, and found DNAmAA estimated by the Horvath clock to be associated with incident MetS diagnosed 10 years later in the CARDIA dataset ^6^. In our study, however, neither the Horvath clock nor any of the other two first-generation clocks tested (Hannum and 7-CpC DNAmAA, Table 4 and Supplementary Table 6) as assessed at baseline were associated with incident MetS at follow-up. It is noteworthy that participants of the CARDIA cohort were significantly younger (40.4 ±3.5 years) than those in BASE-II investigated here (68.2 ±3.5 years), a difference which might have contributed to the slightly discrepant results.

The results of the current study were complemented by ROC statistics indicating a small, but non-significant difference in the AUC predicting MetS from a model including only clinical parameters compared to the use of GrimAge DNAmAA as a single MetS predictor. Interestingly, none of the tested epigenetic biomarkers was able to improve the prediction of incident MetS at follow-up when added to the model with the clinical MetS predictors. This suggests that a large proportion of the informational content of the combined clinical parameters (WHR, systolic blood pressure and HbA1c in combination with age and sex) is already embedded in GrimAge DNAmAA and to a lesser extent in DunedinPACE. While it is beyond the scope of the current study to present a more detailed comparison of the predictive value for incident MetS of more complex and different combinations of clinical parameters and epigenetic age measures our results clearly show that the GrimAge and DunedinPACE estimates are in the same league as the clinical parameters commonly used in risk models predicting the incidence of MetS. The clinical variables are comparatively easy to determine, so that GrimAge and DunedinPACE do not necessarily have an advantage when the main goal is MetS prediction. The potential advantage of using epigenetic biomarkers may become clearer once these markers have been more deeply investigated for other phenotypes. For instance, DunedinPACE has been reported to be associated with increased risk for incident CVD, stroke/TIA, general morbidity & mortality, frailty ^12,34^, and certain blood glycemic traits ^35^ whereas no direct associations with incident type 2 diabetes mellitus have yet described for DunedinPACE ^35,36^. By determining advanced biomarkers such as DunedinPACE in clinical practice, the future risk for a range of phenotypes could be determined in one laboratory measure, whereas the determination of the individual parameters (laboratory and clinical) required for the prediction of these phenotypes would then represent the greater effort.

Moreover, epigenetic clocks were developed with the aim of a more general assessment of an individual’s “biological age” but not for predicting specific diseases. The fact that GrimAge DNAmAA and DunedinPACE still perform similarly well as a simple combination of clinical parameters in the prediction of incident MetS suggests that CpG combinations identified to specifically predict MetS (e.g. as MetS methylation risk scores) would potentially perform even better in the prediction of this outcome.

Our study has several limitations and strengths that we would like to acknowledge. First, due to the exploratory nature of the analyses presented here, we did not correct for multiple testing, so our results must be considered preliminary and need to be confirmed in independent studies. Second, it is well documented that BASE-II participants show an above average health ^13^ which might have impacted our results. The fact that we were still able to detect significant differences suggests that the detected associations may also be present in the general population. Third, and along the same lines, around one third of the original participants could not be re-examined in the follow-up, either due to death or other reasons (e.g. attrition). While a bias due to study dropout is inherent in observational studies in general and was already shown for BASE-II in previous work by our group e.g. with regard to age, education and frailty (Fried), there was no difference in sex distribution and overall morbidity associated with participation in the follow-up assessments ^37,38^. This suggests that the impact of this type of potential bias on our results is likely small.

The main strengths of our study include the comparatively large sample size with an equal representation of men and women, and the availability of comprehensive information on potential confounders allowing to consider them in our analyses. Additionally, the availability of longitudinal data with a comparatively long follow-up period and computation of third generation epigenetic clock algorithms allowed a comprehensive investigation of the association between MetS and epigenetic aging.

In conclusion, our study on the relationship between epigenetic clocks and MetS has confirmed and extended previous results on this topic. A noteworthy extension of our work relates to the direct cross-sectional comparison between epigenetic clock measures of the first, second and third generation within a single dataset, as well as assessments using longitudinal outcome data. These primarily show an association between DunedinPACE and MetS and illustrate the potential use of epigenetic measures for the assessment of disease risk.

## Acknowledgments

This work was supported by grants of the Deutsche Forschungsgemeinschaft (grant number 460683900 to ID and LB), the ERC (as part of the Lifebrain project to LB), and the Cure Alzheimer’s Fund (as part of the CIRCUITS consortium to LB). This article uses data from the Berlin Aging Study II (BASE-II) and the GendAge study which were supported by the German Federal Ministry of Education and Research under grant numbers #01UW0808; #16SV5536K, #16SV5537, #16SV5538, #16SV5837, #01GL1716A and #01GL1716B. J.H. was supported by a grant from the EU Joint Programme – Neurodegenerative Disease Research (JPND2021-650-289, coordinator: C.M.L.). C.M. Lill was supported by the Heisenberg program of the DFG (DFG; LI 2654/4-1). We thank all probands of the BASE-II/GendAge study for their participation in this research.

## Author contributions

Conceived and designed the analyses: ID and VMV. Contributed study specific data: ID, JH, CL and LB. Analyzed the data: ID and VMV. Wrote the manuscript: ID. All authors revised and approved the manuscript.

## Competing interests

The authors declare no competing interests.

## Data availability statement

The data presented in this study are available from the BASE-II office after filling in a data request form that undergoes review, please see https://www.base2.mpg.de/7549/data-documentation for details. We are not in a position to make data publicly available because these contain information that could compromise research participants’ privacy and consent.

---

**Supplementary Figure 1:**
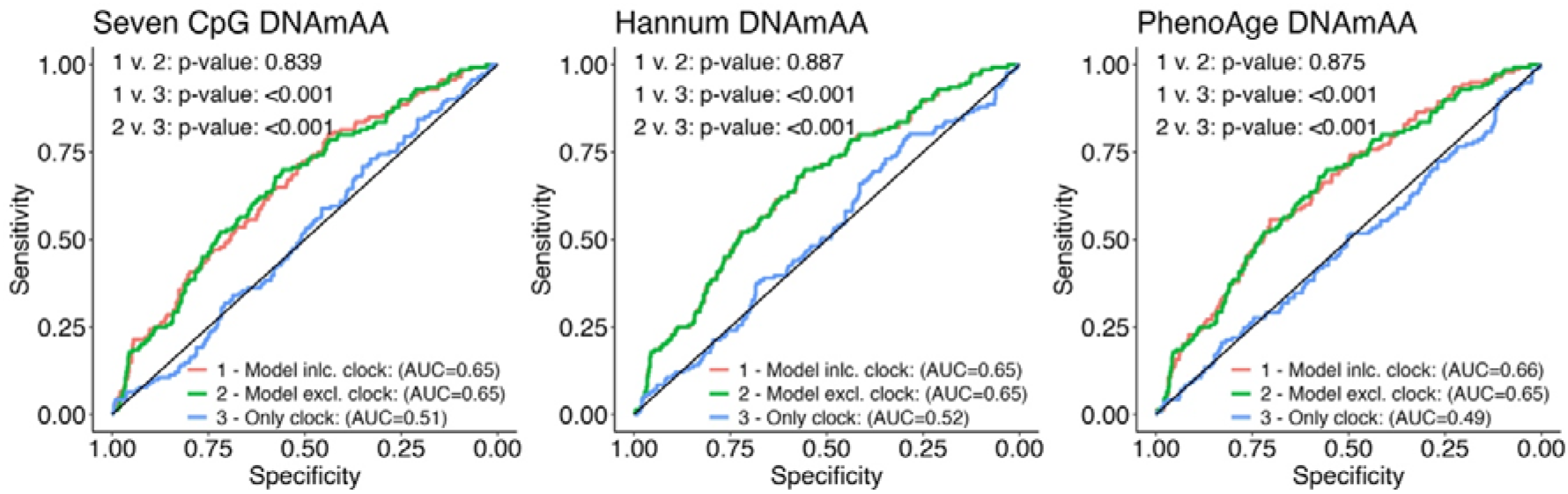
ROC curves derived from logistic regression analyses of incident MetS at follow-up on epigenetic age estimators at baseline (Seven CpG, Hannum and PhenoAge DNAmAA estimates). The red (1) and green (2) curve represent prediction model with and without the epigenetic age estimate. The blue curve (3) visualizes results from a prediction model with only the epigenetic age estimate without any additional variables. The blue curve represents a prediction model that includes only the epigenetic estimate. Note: AUC = Area under the curve, DNAmAA = DNA methylation age acceleration; incl.=including; excl.=excluding.

## Notes

### Competing Interest Statement

The authors have declared no competing interest.

### Author Declarations

All participants gave written informed consent. The medical assessments at baseline and follow-up were conducted in accordance with the Declaration of Helsinki and approved by the Ethics Committee of the Charité - Universitätsmedizin Berlin (approval numbers EA2/029/09 and EA2/144/16).

## References

1. Alberti KG, Eckel RH, Grundy SM, et al. Harmonizing the metabolic syndrome: a joint interim statement of the International Diabetes Federation Task Force on Epidemiology and Prevention; National Heart, Lung, and Blood Institute; American Heart Association; World Heart Federation; International Atherosclerosis Society; and International Association for the Study of Obesity. Circulation. Oct 20 2009;120(16):1640–5. doi:10.1161/CIRCULATIONAHA.109.192644

2. Noubiap JJ, Nansseu JR, Lontchi-Yimagou E, et al. Geographic distribution of metabolic syndrome and its components in the general adult population: A meta-analysis of global data from 28 million individuals. Diabetes Res Clin Pract. Jun 2022;188:109924. doi:10.1016/j.diabres.2022.109924

3. Schutte S, Eberhard S, Burger B, Hemmerling M, Rossol S, Stahmeyer JT. [Prevalence of metabolic syndrome : Analysis based on routine statutory health insurance data]. Inn Med (Heidelb). May 2023;64(5):482–489. Pravalenz des metabolischen Syndroms : Eine Analyse auf Basis von Routinedaten einer gesetzlichen Krankenversicherung. doi:10.1007/s00108-023-01510-4

4. Loh NY, Noordam R, Christodoulides C. Telomere length and metabolic syndrome traits: A Mendelian randomisation study. Aging Cell. Aug 2021;20(8):e13445. doi:10.1111/acel.13445

5. Jansen R, Han LK, Verhoeven JE, et al. An integrative study of five biological clocks in somatic and mental health. Elife. Feb 9 2021;10doi:10.7554/eLife.59479

6. Nannini DR, Joyce BT, Zheng Y, et al. Epigenetic age acceleration and metabolic syndrome in the coronary artery risk development in young adults study. Clin Epigenetics. Nov 15 2019;11(1):160. doi:10.1186/s13148-019-0767-1

7. Horvath S, Raj K. DNA methylation-based biomarkers and the epigenetic clock theory of ageing. Nat Rev Genet. Jun 2018;19(6):371–384. doi:10.1038/s41576-018-0004-3

8. Warner B, Ratner E, Datta A, Lendasse A. A systematic review of phenotypic and epigenetic clocks used for aging and mortality quantification in humans. Aging (Albany NY). Aug 30 2024;16(17):12414–12427. doi:10.18632/aging.206098

9. Lee HS, Park T. The influences of DNA methylation and epigenetic clocks, on metabolic disease, in middle-aged Koreans. Clin Epigenetics. Oct 15 2020;12(1):148. doi:10.1186/s13148-020-00936-z

10. McCarthy K, O’Halloran AM, Fallon P, Kenny RA, McCrory C. Metabolic syndrome accelerates epigenetic ageing in older adults: Findings from The Irish Longitudinal Study on Ageing (TILDA). Exp Gerontol. Nov 2023;183:112314. doi:10.1016/j.exger.2023.112314

11. Fohr T, Hendrix A, Kankaanpaa A, et al. Metabolic syndrome and epigenetic aging: a twin study. Int J Obes (Lond). Jun 2024;48(6):778–787. doi:10.1038/s41366-024-01466-x

12. Belsky DW, Caspi A, Corcoran DL, et al. DunedinPACE, a DNA methylation biomarker of the pace of aging. Elife. Jan 14 2022;11doi:10.7554/eLife.73420

13. Bertram L, Bockenhoff A, Demuth I, et al. Cohort profile: The Berlin Aging Study II (BASE-II). Int J Epidemiol. Jun 2014;43(3):703–12. doi:10.1093/ije/dyt018

14. Gerstorf D, Bertram L, Lindenberger U, et al. Editorial. Gerontology. 2016;62(3):311–5. doi:10.1159/000441495

15. Demuth I, Bertram L, Drewelies J, et al. Berlin Aging Study II (BASE-II). In: Gu D, Dupre ME, eds. Encyclopedia of Gerontology and Population Aging. Springer International Publishing; 2021:649–656.

16. Demuth I, Banszerus V, Drewelies J, et al. Cohort profile: follow-up of a Berlin Aging Study II (BASE-II) subsample as part of the GendAge study. BMJ Open. Jun 23 2021;11(6):e045576. doi:10.1136/bmjopen-2020-045576

17. Horvath S. DNA methylation age of human tissues and cell types. Genome biology. 2013;14(10):R115. doi:10.1186/gb-2013-14-10-r115

18. Hannum G, Guinney J, Zhao L, et al. Genome-wide methylation profiles reveal quantitative views of human aging rates. Molecular cell. Jan 24 2013;49(2):359–367. doi:10.1016/j.molcel.2012.10.016

19. Levine ME, Lu AT, Quach A, et al. An epigenetic biomarker of aging for lifespan and healthspan. Aging (Albany NY). 2018;10(4):573. 10.18632/aging.101414

20. Lu AT, Quach A, Wilson JG, et al. DNA methylation GrimAge strongly predicts lifespan and healthspan. Aging (Albany NY). 2019;11(2):303.

21. Belsky DW, Caspi A, Corcoran DL, et al. DunedinPACE, a DNA methylation biomarker of the pace of aging. eLife. 2022/01/14 2022;11:e73420. doi:10.7554/eLife.73420

22. Sommerer Y, Dobricic V, Schilling M, et al. Epigenome-Wide Association Study in Peripheral Tissues Highlights DNA Methylation Profiles Associated with Episodic Memory Performance in Humans. Biomedicines. Nov 3 2022;10(11)doi:10.3390/biomedicines10112798

23. Vetter VM, Meyer A, Karbasiyan M, Steinhagen-Thiessen E, Hopfenmuller W, Demuth I. Epigenetic clock and relative telomere length represent largely different aspects of aging in the Berlin Aging Study II (BASE-II). The journals of gerontology Series A, Biological sciences and medical sciences. Aug 18 2018;doi:10.1093/gerona/gly184

24. Quach A, Levine ME, Tanaka T, et al. Epigenetic clock analysis of diet, exercise, education, and lifestyle factors. Aging (Albany NY). Feb 14 2017;9(2):419–446. doi:10.18632/aging.101168

25. Horvath S, Gurven M, Levine ME, et al. An epigenetic clock analysis of race/ethnicity, sex, and coronary heart disease. Genome biology. 2016;17(1):1–23.

26. Oblak L, van der Zaag J, Higgins-Chen AT, Levine ME, Boks MP. A systematic review of biological, social and environmental factors associated with epigenetic clock acceleration. Ageing Research Reviews. 2021/08/01/ 2021;69:101348. 10.1016/j.arr.2021.101348

27. Vetter VM, Kalies CH, Sommerer Y, et al. Relationship Between 5 Epigenetic Clocks, Telomere Length, and Functional Capacity Assessed in Older Adults: Cross-Sectional and Longitudinal Analyses. The Journals of Gerontology: Series A. 2022;77(9):1724–1733. doi:10.1093/gerona/glab381

28. Meyer A, Salewsky B, Spira D, Steinhagen-Thiessen E, Norman K, Demuth I. Leukocyte telomere length is related to appendicular lean mass: cross-sectional data from the Berlin Aging Study II (BASE-II). Am J Clin Nutr. Jan 2016;103(1):178–83. doi:10.3945/ajcn.115.116806

29. Charlson ME, Pompei P, Ales KL, MacKenzie CR. A new method of classifying prognostic comorbidity in longitudinal studies: development and validation. J Chronic Dis. 1987;40(5):373–83. doi:10.1016/0021-9681(87)90171-8

30. Nothlings U, Hoffmann K, Bergmann MM, Boeing H. Fitting portion sizes in a self-administered food frequency questionnaire. J Nutr. Dec 2007;137(12):2781–6. doi:10.1093/jn/137.12.2781

31. Tan C, Sasagawa Y, Kamo KI, et al. Evaluation of the Japanese Metabolic Syndrome Risk Score (JAMRISC): a newly developed questionnaire used as a screening tool for diagnosing metabolic syndrome and insulin resistance in Japan. Environ Health Prev Med. Nov 2016;21(6):470–479. doi:10.1007/s12199-016-0568-5

32. R Core Team (2018). R: A language and environment for statistical computing. R Foundation for Statistical Computing, Vienna, Austria. Updated 16.10.2024. https://www.R-project.org/

33. Vetter VM, Demircan K, Homann J, et al. Low Blood Levels of Selenium, Selenoprotein P and GPx3 are Associated with Accelerated Biological Aging: Results from the Berlin Aging Study II (BASE-II). medRxiv. 2024:2024.04. 04.24305314.

34. Mak JKL, Karlsson IK, Tang B, et al. Temporal Dynamics of Epigenetic Aging and Frailty From Midlife to Old Age. The journals of gerontology Series A, Biological sciences and medical sciences. Oct 1 2024;79(10)doi:10.1093/gerona/glad251

35. Miao K, Hong X, Cao W, et al. Association between epigenetic age and type 2 diabetes mellitus or glycemic traits: A longitudinal twin study. Aging Cell. Jul 2024;23(7):e14175. doi:10.1111/acel.14175

36. Wikstrom Shemer D, Mostafaei S, Tang B, et al. Associations between epigenetic aging and diabetes mellitus in a Swedish longitudinal study. Geroscience. Oct 2024;46(5):5003–5014. doi:10.1007/s11357-024-01252-7

37. Spieker J, Vetter VM, Drewelies J, et al. Diabetes type 2 in the Berlin Aging Study II: Cross-sectional and longitudinal data on prevalence, incidence and severity over on average seven years of follow-up. Diabet Med. Aug 2023;40(8):e15104. doi:10.1111/dme.15104

38. König M, Malsch C, Mariño J, et al. Nocturia as a Risk Factor for Developing Frailty in Older Adults: Results of the Berlin Aging Study II. medRxiv. 2024:2024.09.20.24313292. doi:10.1101/2024.09.20.24313292

