## Supplementary Methods for "DunedinPACE Predicts Incident Metabolic Syndrome: Cross-sectional and Longitudinal Data from the Berlin Aging Study II (BASE-II)"

Potential participants were recruited via a participant pool at the Max Planck Institute for Human Development (Berlin, Germany) and via advertisements placed in local newspapers and the public transportation system. Participants between the ages of 60 and 80 (older group) were eligible for recruitment.

A comprehensive medical follow-up including 1,083 participants of the older group took place between 2018 and 2020 after an average follow-up period of 7.4 (±1.5 SD) years [1]. Seventeen additional participants of the older age group were assessed at follow-up examination only.

All participants gave written informed consent. The medical assessments at baseline and follow-up were conducted in accordance with the Declaration of Helsinki and approved by the Ethics Committee of the Charité – Universitätsmedizin Berlin (approval numbers EA2/029/09 and EA2/144/16) and were registered in the German Clinical Trials Registry as DRKS00009277 and DRKS00016157.

*Estimators of epigenetic age and DunedinPACE*

Outliers in the DNAm fractions were identified using the *outlyx* and *pcout* function from the "bigmelon"-package [2]. Samples with a bisulfite conversion efficiency below 80% were excluded from the dataset. The resulting sample set was normalized using the *dasen* function. Samples that showed a difference of a root-mean squared deviation of 0.1 or more in beta-values after normalization compared to the original dataset were excluded from subsequent analyses. The final estimations of Horvath's, Hannum's, PhenoAge, and GrimAge DNAmA were computed using the DNA Methylation Age Calculator (https://dnamage.clockfoundation.org/) using default parameters. DunedinPACE was calculated using the PACEProjector() function from the DunedinPACE R package (https://github.com/danbelsky/DunedinPACE). The 7-CpG clock was trained in baseline samples of the BASE-II and was validated in two independent cohorts [3,4] and in context of a number of phenotypes [5-10]. It is calculated from methylation data measured using methylation-sensitive single-nucleotide primer extension (MS-SNuPE) [11]. A detailed protocol as well as additional information about the comparability of the SNuPE method with results from the Illumina array were described in detail before [5,8,12].

### References


1. Demuth I, Banszerus V, Drewelies J, et al. Cohort profile: follow-up of a Berlin Aging Study II (BASE-II) subsample as part of the GendAge study. *BMJ Open*. Jun 23 2021;11(6):e045576. doi:10.1136/bmjopen-2020-045576
2. Gorrie-Stone TJ, Smart MC, Saffari A, et al. Bigmelon: tools for analysing large DNA methylation datasets. *Bioinformatics*. 2019;35(6):981-986.
3. Banszerus VL, Vetter VM, Salewsky B, König M, Demuth I. Exploring the relationship of relative telomere length and the epigenetic clock in the LipidCardio cohort. *International journal of molecular sciences*. 2019;20(12):3032. doi:10.3390/ijms20123032
4. Feldkamp JD, Vetter VM, Arends CM, et al. CHIP-related epigenetic age acceleration correlates with CHIP clone size in patients with high morbidity. *Haematologica*. 03/03 2022;doi:https://doi.org/10.3324/haematol.2021.280021
5. Vetter VM, Meyer A, Karbasiyan M, Steinhagen-Thiessen E, Hopfenmuller W, Demuth I. Epigenetic clock and relative telomere length represent largely different aspects of aging in the Berlin Aging Study II (BASE-II). *The journals of gerontology Series A, Biological sciences and medical sciences*. Aug 18 2018;doi:10.1093/gerona/gly184
6. Vetter VM, Demircan K, Homann J, et al. Low Blood Levels of Selenium, Selenoprotein P and GPx3 are Associated with Accelerated Biological Aging: Results from the Berlin Aging Study II (BASE-II). *medRxiv*. 2024:2024.04. 04.24305314.
7. Vetter VM, Drewelies J, Sommerer Y, et al. Epigenetic aging and perceived psychological stress in old age. *Translational Psychiatry*. 2022/09/26 2022;12(1):410. doi:https://doi.org/10.1038/s41398-022-02181-9
8. Vetter VM, Kalies CH, Sommerer Y, et al. Relationship Between 5 Epigenetic Clocks, Telomere Length, and Functional Capacity Assessed in Older Adults: Cross-Sectional and Longitudinal Analyses. *The Journals of Gerontology: Series A*. 2022;77(9):1724-1733. doi:10.1093/gerona/glab381
9. Vetter VM, Sommerer Y, Kalies CH, Spira D, Bertram L, Demuth I. Vitamin D supplementation is associated with slower epigenetic aging. *GeroScience*. 2022/05/13 2022;doi:https://doi.org/10.1007/s11357-022-00581-9
10. Vetter VM, Spieker J, Sommerer Y, et al. DNA methylation age acceleration is associated with risk of diabetes complications. *Communications Medicine*. 2023;3(1):21.
11. Kaminsky ZA, Assadzadeh A, Flanagan J, Petronis A. Single nucleotide extension technology for quantitative site-specific evaluation of metC/C in GC-rich regions. *Nucleic acids research*. Jun 15 2005;33(10):e95. doi:10.1093/nar/gni094
12. Vetter VM, Kalies CH, Sommerer Y, Bertram L, Demuth I. Seven-CpG DNA Methylation Age Determined by Single Nucleotide Primer Extension and Illumina's Infinium MethylationEPIC Array Provide Highly Comparable Results. Original Research. *Frontiers in Genetics*. 2022-January-17 2022;12doi:https://doi.org/10.3389/fgene.2021.759357
